# Approaching Patient-Clinician Relationships Issues Involving Artificial Intelligence Using Ethics of Care and Nursing Ethics

**DOI:** 10.1101/2020.04.21.20073544

**Authors:** Soaad Q. Hossain

## Abstract

With the rise of artificial intelligence (AI) and its application within industries, there is no doubt that someday AI will be one of the key players in medical diagnoses, assessments and treatments. With the involvement of AI in health care and medicine comes concerns pertaining to its application, more specifically its impact on both patients and medical professionals. To further expand on the discussion, using ethics of care, literature and a systematic review, we will address the impact of allowing AI to guide clinicians with medical procedures and decisions. We will then argue that the impact of allowing AI to guide clinicians with medical procedures and decisions can hinder patient-clinician relationships, concluding with a discussion on the future of patient care and how ethics of care can be used to investigate issues within AI in medicine.

## Introduction

As artificial intelligence (AI) becomes accessible to medicine, there is no doubt that someday it will be a key player in medical diagnoses, assessments and treatments. Consequently, there are concerns pertaining to its application, more specifically its impact on both patients and clinicians. Using ethics of care and nursing ethics literature, we will argue that the impact of allowing AI to guide clinicians with medical procedures and decisions can hinder patient-clinician relationships. We will first define AI and ethics of care, then review literature on the role of AI in medicine, and ethics of care. Then, we will conduct a systematic review on moral distress in healthcare professionals and clinician-nurse relationships. Accordingly, we will describe the findings and use the results to evaluate the impact of allowing AI to guide clinicians with medical procedures and decisions. We will conclude with a discussion on the future of patient can and emphasize how ethics of care can be used to investigate issues in AI and medicine.

## Preliminaries

### Artificial Intelligence

We define AI as the scientific discipline that aims to understand and develop systems that display properties of intelligence (Panch, Szolovits & Atun, 2018). The role of AI in medicine would be the following: laboratory information (e.g. analyze patient data for infections and breakouts), time reduction and optimization (i.e. reducing time of health care and medical procedures), medical decision support (e.g. assisting with deciding treatment plans), triage and screening tool, and interpretation, annotation and comprehension of health care data.

### Ethics of Care

Also known as care ethics, ethics of care can be understood as the effort and the art of understanding a person in their reality of suffering (Schuchter & Heller, 2017). That is, it is the effort and art of entering with others into compassionate and caring relations. What makes ethics of care important is that it has the potential of being based on the truly universal experience of care (Held, 2006). As Virginia Held mentions in her book *The Ethics of Care – Personal, Political and Global*, ethics of care develops based on experience, reflection on it and discourse concerning it (Held, 2006). Accordingly, the use of it enables us to better understand problems associated with care.

## Literature Review

### Artificial Intelligence in Medicine

Given that AI can perform certain tasks with greater consistency, speed, and reproducibility than humans (He et al., 2019), it is said that AI can assist clinicians in shortening processing times and improve the quality of patient care in clinical practice (Krittanawong, 2018). For instance, AI can be used for medical decision support, and as a screening tool. AI can automate some of the complex and time-consuming tasks involved in pathology such as object quantification and rare target identification (Mason, Morrison & Visintini, 2018). However, despite its capabilities, several concerns were raised within the literature; specifically, its effects on clinicians and patients. A concern expressed for patients is that with the application of AI in medicine, there is a potential for an increased feeling of objectification and the loss of control as well as deception and infantilization within patients, especially for the elderly (Becker, 2019). The concern relates to clinician-patient relationship, which is that for patients, there a reduction in the amount of human contact (Becker, 2019). Also, a concern is that AI applications (e.g. those that monitor speech and behavior) could change the relationship between clinicians and patients by generating fear that data collected could be scrutinized and used for health care decisions (Mason, Morrison & Visintini, 2018); and that the issue of trust when both clinicians and patients discuss and accept the recommendations provided by AI (Noorbakhsh-Sabet et al., 2019).

### Ethics of Care and Nursing Ethics

Ethics of care is primarily response-focused; focuses on the fundamental dependence of individuals and using relationships to view ethical problems. Within relationships, ethics of care investigates the reason or justification for a particular action or reason (Krause & Boldt, 2018). As a result, what fields such as nursing ethics has done was use it to inquire about ethical issues in care. In utilizing it, this led to recent literature concluding that ethics of care is an interdisciplinary field of inquiry which is driven by social concerns (Leget, Nistelrooji & Visse, 2019). Additionally, what was discovered was issues surround medical practice, values and morality.

Literature on nursing ethics and ethics of care have expressed concerns of the values essential to the moral foundation of nursing cannot be extracted from any abstract or decontextualized moral theory (Parker, 1990). These concerns have expanded to concerns surrounding moral distress, with an increasing number of published studies reporting moral distress as a common problem among healthcare professionals (Sasso et al., 2016). Moral distress is the psychological, emotional and psychological suffering that may be experienced when one acts in a way that is not consistent with one‚s deeply held ethical values, principles or moral commitments (Sasso et al., 2016). Investigating moral distress led to several bioethical issues. In the case of emergency care, nurses have reported ethical conflicts in emergency situations when they and the physician(s) did not share the same paradigm, and when the nurse felt that they were in the middle between the patient and the physician (Jimenez-Herrera & Axelsson, 2015). Accordingly, they state that those situations usually occur when the solution for the patient was only based on a biomedical perspective but required a more integrative model from a holistic perspective (Jimenez-Herrera & Axelsson, 2015). Furthermore, nurses felt that the patient, assisted in the critical situation often are reified and fragmented to a number or a disease, leading to depersonalized and dehumanized caring processes (Jimenez-Herrera & Axelsson, 2015).

## Methods

### Search Outcome and Quality Appraisal

We systematically reviewed articles that study the root cause of moral distress in clinicians and an article that studied clinician-nurse relationships. We conducted an online search on Google Scholar for both types of studies. The keywords that we used in our search are “moral distress”, “physicians”, “nurses”, and “clinicians”. We then used the following set of criteria to select the moral distress articles:

1. The article must discuss about moral distress experienced by physicians, nurses and/or direct or indirect care providers.
2. The article must contain quantitative results on the root cause of moral distress within nurses and/or direct or indirect care providers

We used the following criteria to select the physician-nurse relationship article:

1. The article must discuss about issues experienced by physicians and nurses when working together.
2. The article must contain qualitative results on physician-nurse relationships.

Conducting the search led us to obtaining 4 relevant articles. Using the results from the 4 articles concurrently with ethics of care, we perform the ethical analysis to evaluate the impact of allowing AI to guide clinicians with medical procedures and decisions.

### Methodology and Data Analysis

The first three articles pertain to moral distress. The fourth article pertains to physician-nurse relationships. The first article by Whitehead et al. conducted a web-based survey of demographics, Moral Distress Scale-Revised (MDS-R) and a short version of Olson‚s Hospital Ethical Climate Scale (HECS-S) to gather data. The second article by Lusignani et al. conducted a cross-sectional questionnaire survey to evaluate the frequency, intensity and levels of moral destress. The third article by Epstein et al. used data from previous studies, write-in items, moral distress consultations and recent publications from professionals in which root causes of moral distress were described to revise the MDS-R to then test the Measure of Moral Distress for Healthcare professionals (MMD-HP). The fourth article by Sabone et al. conducted semi-structured interviews at public hospitals.

## Results

### Findings

The studies on the root cause of moral distress in clinicians with respect to the study on the physician-nurse relationship show that there are three trends. The first trend is the communication issues between the physician and the nurse or others care providers (i.e. direct care or indirect care providers). The second trend is the diminished or lack of care for the patient from the physicians and the nurses. The third trend is the patient‚s family wishing to continuing treatment of the patient‚s when it is not in the best interest of the patient. Figure 1 displays the findings from the studies on the root cause of moral distress in clinicians while figure 2 displays the study on the physician-nurse relationship.

**Figure 1:**
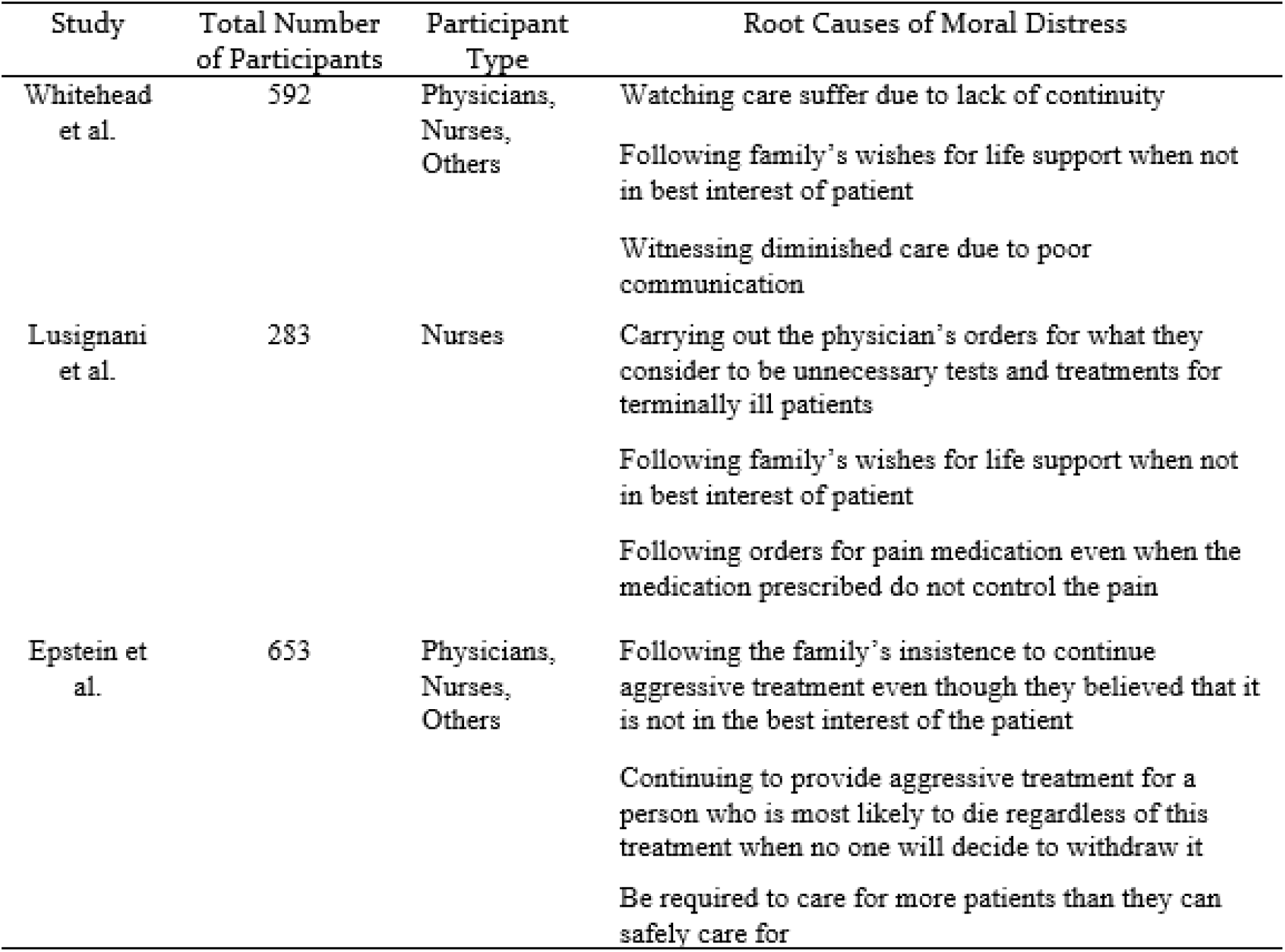
Table displaying the findings from the studies on the root causes of moral distress. Others in participant type are direct or indirect care providers. The root causes of moral distress listed are the top three causes mentioned within each study, respectively.

**Figure 2:**
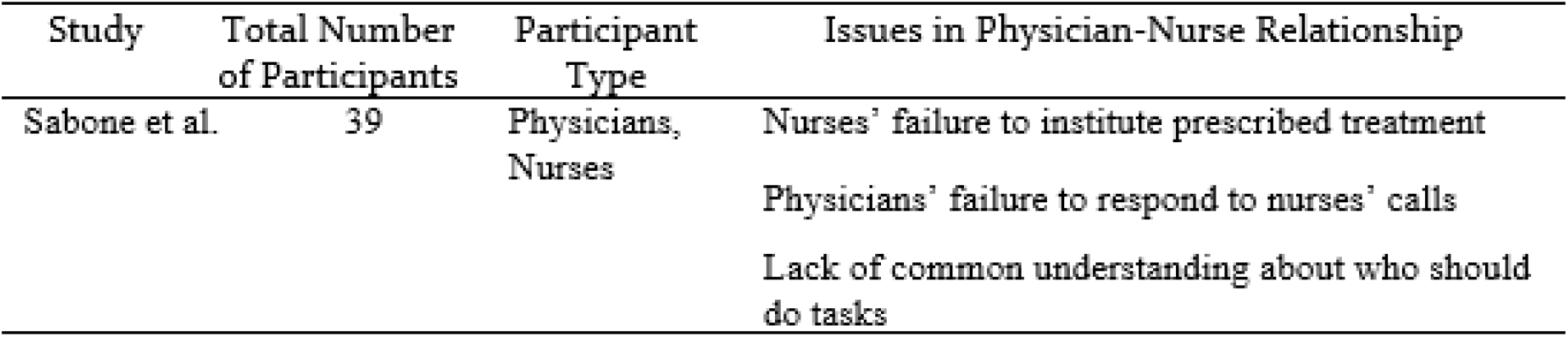
Table displaying the findings from the study on physician-nurse relationships, displaying the top three issues that contribute to strained physician-nurse relationships.

### Physician-Nurse Relationships

Given the first trend, the impact of allowing AI to guide clinicians with medical procedures and decisions is that it can worsen the communication issues between the physician and the nurses. Given that AI‚s high accuracy and success in performing medical tasks and its wide range of application in medicine, this will motivate clinicians to follow through with the guidance provided by AI. As such, clinicians will then utilize and listen to AI to assist them in medical procedures and decisions over nurses for two reasons. The first being that in addition to AI‚s performance, current AI is designed to share the same interest and paradigm as clinicians when it comes to how they approach health care. AI is focused on successfully treating the patient, not caring for it. Accordingly, when AI guides clinicians, it would guide it in a way that agrees with the interest of clinicians. When clinicians realize this, this will encourage them to listen to AI more often, eventually reaching a point where they will listen to AI over nurses for two reasons

The second reason is due to the existing communication issue between clinicians and nurses. The results found within the studies on the root cause of moral distress and physician-nurse relationship highlight that there is a lack of communication between the physician and the nurse. In physicians having different interest and paradigm than that of nurses, these negatively impact the communication between physicians and nurses by having physicians focus their effort on communicating medical procedures and decisions to nurses than having discussions on that and patient care with them. With AI involved, this will worsen the communication by reinforcing physicians‚ interest and paradigm, further enabling physicians to focus their effort on communicating medical procedures and decisions to nurses. Consequently, occurrences of moral distress within clinicians and nurses increase.

### Patient-Clinician Relationships

Given the second trend, the impact of allowing AI to guide clinicians with medical procedures and decisions is that it can further diminish the quality of care for the patients from the physicians and the nurses. With current AI being designed to have the same interest and paradigm as clinicians, this enables AI to easily create a bond with clinicians. As the bond between AI and clinicians strengthen, this would decrease the bond between clinicians and patients. The reason for this occurrence would be because the relationship between clinicians and AI would become better than that of clinicians and patients as different from patients, both AI and clinicians share the same interest and paradigm. Consequently, this promotes the reduction in the amount of human contact within patient-clinician relationships. What results from this is increased feelings of objectification and loss of control as well as deception and infantilization within patients.

## Discussion

### Future of Patient Care

The impact of allowing AI to guide clinicians with medical procedures and decisions can ultimately lead to creating or fortifying depersonalized and dehumanized caring processes. With clinicians applying AI, the solution for patients will continue to be only based on a biomedical perspective and not a more integrative model from a holistic perspective. Accordingly, the patients being assisted in the critical situation will increasingly become reified and fragmented to a number or a disease, leading to depersonalized and dehumanized caring processes.

## Conclusion

Through using ethics of care to investigate ethical issues in AI in medicine, we found that the concerns brought up in the literature on AI and nursing ethics are not only valid concerns, but those concerns are at risk of becoming realized or more problematic. With the use of AI in medicine, we can forecast an increase in moral distress within health care professionals, diminish clinician-nurse relationships, and diminish patient-clinician relationships.

## Data Availability

Data was taken from 4 studies. Links to the studies can be found below.
Study 1: https://www.ncbi.nlm.nih.gov/pubmed/25440758
study 2: https://www.ncbi.nlm.nih.gov/pubmed/31002584
Study 3: https://www.ncbi.nlm.nih.gov/pubmed/27726233s
Study 4: https://www.ncbi.nlm.nih.gov/pubmed/31014168

## References

Becker A. (2019). Artificial Intelligence in Medicine: What is it Doing for us Today? Elsevier Inc. 8. 200–201.

Brody H, & Clark M. (2014). Narrative Ethics: A Narrative. Hastings Center Report.

Epstein E. G., Whitehead P. B., Prompahakul C., Thacker L. R., & Hamric A. B. 2019. Enhancing Understanding of Moral Distress: The Measure of Moral Distress of Health Care Professionals. AJOB Empirical Bioethics. 10. 2. 113–124.

Franziska K., & Boldt J. (2018). Care in Healthcare – Reflections on Theory and Practice. Palgrave Macmillan. 55–62.

He J., Baxter S. L., Xu Jie., Xu Jim., Zhou X. & Zhang K. (2019). The Practical Implementation of Artificial Intelligence Technologies in Medicine. Nature Medicine. 25. 30–35.

Held, V. (2006). The Ethics of Care – Personal, Political and Global. Oxford University Press. 3.

Jimenez-Herrera M. F., & Axelsson C. (2015). Some Ethical Conflicts in Emergency Care. Sage. 22. 5. 551–553.

Krittanawong C. (2018). The Rise of Artificial Intelligence and the Uncertain Future for Physicians. European Journal of Internal Medicine. 48. 13–14.

Leget C., Nistelrooji I. V., & Visse M. (2019). Beyond Demarcation: Care Ethics as an Interdisciplinary Field of Inquiry. Sage. 26. 1. 21.

Lusignani M., Gianni M. L., Re L. G., & Buffon M. L. 2017. Moral Distress Among Nurses in Medical, Surgical and Intensive-Care Units. Journal of Nursing Management. 25. 477–485.

Martindale A., & Fisher P. (2019). Disrupted Faces, Disrupted Identities? Embodiment, Life Stories and Acquired Facial ‘Disfigurement’. Foundation for the Sociology of Health & Illness. 20. 1.

Mason J., Morrison A., & Visintini S. (2018). CADTH Issues in Emerging Health Technologies – Informing Decisions about New Health Technologies. CADTH. 174. 7 –12.

Mitchell M. (2019). Artificial Intelligence Hits the Barrier of Meaning. Information. 10. 51. 2.

Noorbakkhsh-Sabet N., Zand R., Zhang Y., & Abedi V. (2019). Artificial Intelligence Transforms the Future of Health Care. Elsevier Inc. 000. 000. 3–5.

Panch T., Szolovits P., & Atun R. (2018). Artificial Intelligence, Machine Learning and Health Systems. Journal of Global Health. 8. 2. 2.

Parker R. S. (1990). Nurses’ Stories. The Search for Relational Ethic of Care. Aspen Publishers Inc. 13. 1. 35.

Sabone M., Mazonde P., Cainelli F., Maitshoko M., Joseph R., Shayo J., Morris B., Muecke M., Wall B. M., Hoke L., Peng L., Mooney-Doyle K., & Ulrich C. M. 2020. Everyday Ethical Challenges of Nurse-Physician Collaboration. Nursing Ethics. 27. 1. 206–220.

Sasso L., Bagnasco A., Bianchi M, Bressan V., & Carneval F. (2016). Moral Distress in Undergraduate Nursing Students: A Systematic Review. Sage. 23. 5. 524–525.

Schuchter P., & Heller A. (2017). The Care Dialog: The “Ethics of Care” Approach and its Importance for Clinical Ethics Consultations. Medicine, Health Care, and Philosophy. 21. 1. 55.

Wahl, B., Cossy-Gantner A., Germann S. & Schwalbe (2018). Artificial Intelligence (AI) and Global Health: How can AI Contribute to Health in Resource-Poor Settings? BMJ Global Health. 3. 1–3.

Whitehead P. B., Herbertson R. K., Hamric A. B., Epstein E. G., & Fisher J. M. 2015. Moral Distress Among Healthcare Professionals: Report of an Institution-Wide Survey. Journal of Nursing Scholarship. 47. 2. 117–125.

